# COVID-19 Symptoms and Immunotherapy in People with Multiple Sclerosis: An Analysis of the COVID-19 in MS Global Data Sharing Initiative Dataset

**DOI:** 10.1101/2023.08.23.23294509

**Authors:** Maria A. Garcia-Dominguez, Bahadar S. Srichawla, Vincent Kipkorir

**Affiliations:** Department of Neurology, University of Massachusetts Chan Medical School; Department of Medicine, University of Nairobi

**Keywords:** COVID-19, multiple sclerosis, intensive care unit, disease-modifying therapy, people with multiple sclerosis, SARS-CoV-2, hospitalization

## Abstract

**OBJECTIVES:** To analyze the symptoms and severity of coronavirus disease 2019 (COVID-19) in people with multiple sclerosis (pwMS) on immunotherapy using data from the COVID-19 in multiple sclerosis (MS) Global Data Sharing Initiative dataset provided by PhysioNet.

**METHODS:** The open-access COVID-19 in MS Global Data Sharing Initiative dataset was obtained through credentialed access using PhysioNet. The variables analyzed included body mass index (BMI), symptoms of COVID-19, age, current use of disease-modifying therapy (DMT), efficacy of DMT, comorbidities, hospitalization status, and type of MS. A linear regression analysis was completed. Data analysis and visualization were completed using STATA *v1.5*, R-Studio *v1.1.447*, Python *v3.8*, and its associated libraries, including NumPy, Pandas, and Matplotlib.

**RESULTS:** A total of 1141 participants were included in the analysis. 904 women and 237 men were diagnosed with MS. Among the pwMS included in the study; 208 (19.54%) had a suspected infection with COVID-19 and only 49 (5.25%) were confirmed. Any COVID-19 symptom was present in 360 individuals. The commonly reported DMT agents included dimethyl fumarate (12.71%) and fingolimod (10.17%). 101 in total (8.85%) reported not using any DMT. Factors associated with hospitalization and/or admission to the ICU included having any comorbidity (*p = 0.01*), neuromuscular disorder (*p = 0.046*), hypertension (*p = 0.005)*, chronic kidney disease (*p < 0.001)*, and immunodeficiency (*p = 0.003*). The type of MS, the duration of the disease, and high-efficacy DMT therapy did not have a statistically significant influence on hospitalization.

**CONCLUSION:** This study underscores the importance of comorbidities, especially neuromuscular disorders, hypertension, chronic kidney disease (CKD), and immunodeficiencies, as possible prognostic indicators for worse outcomes of COVID-19 in pwMS. On the contrary, the type of MS, the duration of the disease, and the efficacy of disease-modifying therapy did not significantly affect the severity of the symptoms of COVID-19 in this cohort.

## INTRODUCTION

The emergence of the novel coronavirus disease 2019 (COVID-19), caused by the severe acute respiratory syndrome coronavirus 2 (SARS-CoV-2), has resulted in a global pandemic with significant morbidity and mortality^1^. As the pandemic has evolved, researchers have aimed to identify specific populations that may be at increased risk of adverse outcomes from the virus. One of such populations of interest is people with multiple sclerosis (pwMS).

Multiple sclerosis (MS) is a chronic autoimmune disease characterized by inflammation, demyelination, and neurodegeneration in the central nervous system ^2^. MS patients often require immunosuppressive or immunomodulatory treatments, which can alter their susceptibility to infections, including viruses such as SARS-CoV-2. Initial concerns arose that these patients might be at higher risk of severe COVID-19 outcomes due to their underlying autoimmune disease and the disease-modifying therapies (DMTs) they receive ^3^. However, the exact relationship between MS and COVID-19 outcomes remains to be fully elucidated. Some studies suggest that the overall risk for people with MS contracting COVID-19 does not appear to increase, but once infected, the course and outcomes of the disease can vary ^4^.

With the availability of large-scale open-access databases, such as the MS Global Data Sharing Initiative dataset from PhysioNet, a comprehensive analysis can provide valuable insights into the interplay between MS and COVID-19. PhysioNet is an open-access resource dedicated to the collaborative development and sharing of biomedical datasets and software. Established by the National Institute of Biomedical Imaging and Bioengineering (NIBIB) and the National Institute of General Medical Sciences (NIGMS), it serves as a platform for researchers worldwide to share and study complex biomedical and physiological data ^5^.

In this study, our objective was to explore the effects of immunomodulatory therapies on COVID-19 in pwMS using an open-access database from the MS Global Data Sharing Initiative, offering a robust perspective on the implications of this global health crisis and providing guidance on future viral pandemics in pwMS ^6^.

## METHODS

### Data Source & Characteristics

This open-access data set was obtained from PhysioNet (*https://physionet.org/content/patient-level-data-covid-ms/1.0.0/*) ^6^. The data collection methodology has been previously described by *Peeters et al*. ^*7*^. Briefly, the data set was collected through a data entry tool that allowed clinicians, pwMS, or their healthcare agents to enter data directly into a central platform of the COVID-19 and MS Global Data Sharing Initiative. The tool was taken down on February 2nd, 2022. Numerous variables were recorded, including patient demographics, COVID-19 symptoms, hospitalization, admission to the intensive care unit (ICU), DMTs, and expanded disability status scale (EDSS). Researchers who accessed the anonymized database completed the required courses to obtain credentialed access. The requirement of individual patient consent was waived because the project did not directly access patient records. The following COVID-19 outcomes are defined. Level 0: If the person has COVID-19 but has not been hospitalized. Level 1: The person has COVID-19 and has been hospitalized. Level 2: The person has COVID-19, has been hospitalized, has been in the intensive care unit, and/or was in a ventilation facility. No individuals within this data set died from COVID-19.

Disease Modifying Therapies (DMTs) for multiple sclerosis (MS) are categorized according to their efficacy in reducing relapse rates, preventing disability progression, and managing magnetic resonance imaging (MRI) activity (new or enlarging lesions). Their classification into low, medium, or high efficacy is often based on a combination of clinical trial data, real-world evidence, and their mechanism of action. The DMT variable was classified as low, medium, and high efficacy. Low efficacy included glatiramer, interferon, and teriflunomide; medium efficacy included fingolimod, cladribine, and dimethyl fumarate; high efficacy included ocrelizumab, alemtuzumab, natalizumab, and rituximab. A category for other categories was included if not listed. This study was completed according to the Strengthening the Reporting of Cohort Studies in Surgery (STROCSS) criteria ^8^.

### Statistical analysis

Descriptive statistics were used to summarize clinical data. Categorical variables were summarized with frequencies. Categorical variables were compared between category etiology groups using Fisher’s exact test. Association between the COVID-19 outcomes and the following variables: The type of MS, duration of the disease, admission to the ICU, being overweight, age, currently on a DMT, DMT efficacy, never treated with a DMT, and comorbidities were evaluated using a univariate linear regression model. All analyses were performed using STATA/IC *ver 15*, R-Studio *ver 1.1.447*, Python *ver 3.8* including the statistical packages Pandas *ver 1.3.2* and NumPy 1.19. Data visualization was completed using the Python library Matplotlib *ver 3.4.3*. A p-value of less than 0.05 was considered statistically significant.

## RESULTS

A total of 1141 participants, 904 women and 237 men, were diagnosed with multiple sclerosis. 883 were in age category 1 (include age range) and 258 in age category 2 (include age range). Among the multiple sclerosis patients included in the study, 858 had no suspected COVID-19, 208 patients had a suspected COVID-19 infection, and 49 were confirmed. A total of 360 participants reported any symptoms of COVID-19. 101 reported chills, 196 dry coughs, 239 fatigue, 141 fever, 88 loss of smell and taste, 156 nasal congestion, 186 pain, 14 pneumonia, 103 shortness of breath, and 169 sore throat.

The DMT most frequently reported was an unspecified drug not listed in the dataset, with 185 occurrences (16.21%). This was closely followed by dimethyl fumarate with 145 instances (12.71%) and fingolimod with 116 occurrences (10.17%). A notable number of participants, 101 in total (8.85%), reported not using any DMT. Other therapies such as interferon, reported by 94 participants (8.24%), ocrelizumab with 84 occurrences (7.36%), and natalizumab, reported by 65 individuals (5.70%), were also substantially represented in the data. The less-used DMT included glatiramer with 64 occurrences (5.61%), teriflunomide with 63 instances (5.52%), and cladribine, chosen by 35 participants (3.07%). The least represented DMTs in the study were rituximab with 15 occurrences (1.31%) and alemtuzumab with 13 instances (1.14%). Additionally, data were not available for 161 individuals, which were excluded from percentage calculations (**Figure 1**).

**Figure 1:**
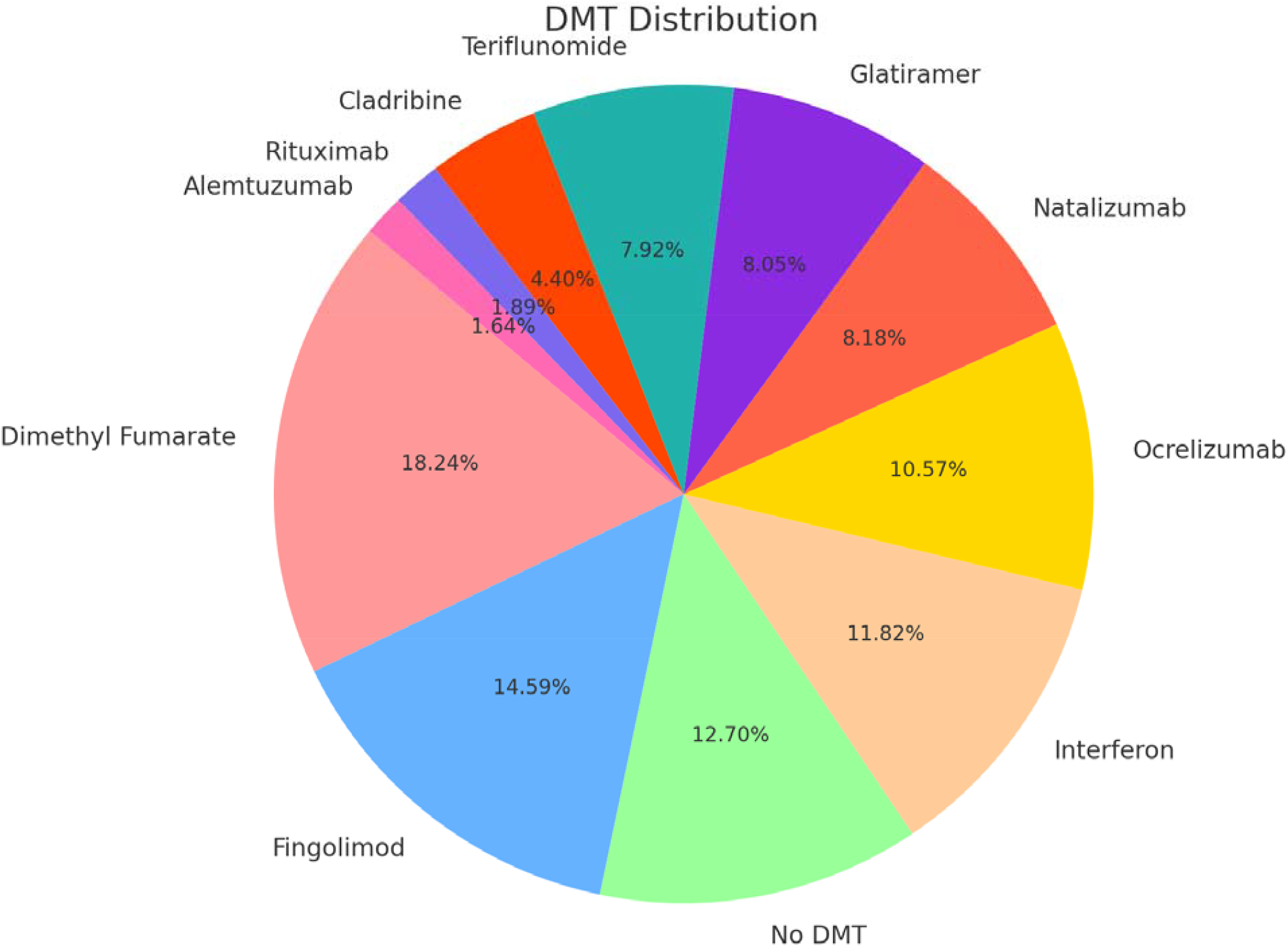
Distribution of disease-modifying therapies (DMTs) among people with multiple sclerosis.

Four patients were admitted to the ICU and required ventilation. 880 were currently on DMT, and 48 were on corticosteroids. A total of 148 individuals reported having at least one comorbidity. Factors associated with worse COVID-19-related outcomes include admission to the ICU (*B 7.33*; *p < 0.0001*), having any comorbidity (*B 1.02*; *p = 0.017*), hypertension (*B 1.11*; *p = 0.005*), chronic kidney disease (*B 1.62; p < 0.0001*) neuromuscular disorder (*B 1.07; p = 0.046*), and immunodeficiency (*B 1.09*; *p = 0.03*). The type of MS (e.g., relapsing-remitting, primary progressive, etc.), duration of the disease, and the DMT efficacy group did not have a statistically significant influence on the severity of COVID-19 symptoms (**Table 1**).

**Table 1:** Variables analyzed, and corresponding p-value based on COVID-19 outcomes; level 0: non-hospitalized and level 1 and 2: hospitalized. *: Fisher exact test CKD: Chronic kidney disease. DM: Diabetes mellitus. DMT: Disease-modifying therapy. MS: Multiple sclerosis. RR: Relapsing remitting.

| Variable(s) | COVID-19<br>Outcomes 0 | COVID-19<br>Outcomes 1 & 2 | P value |
| --- | --- | --- | --- |
| Gender |  |  |  |
| Female | 890 | 14 | 0.33* |
| Male | 236 | 1 |  |
| Age |  |  |  |
| 1 | 883 | 10 | 0.35* |
| 2 | 253 | 5 |  |
| BMI |  |  |  |
| <25 | 852 | 4 | 0.17* |
| >25 | 33 | 1 |  |
| Confirmed case | 49 | 11 | **0.0001** |
| COVID-19 |  |  |  |
| symptoms | 135 | 6 | 0.057 |
| Fever | 88 | 0 | 0.163 |
| Loss of smell/taste | 11 | 3 | 0.003 |
| Pneumonia |  |  |  |
| Required ventilation | 0 | 4 | **0.001** |
| Current DMT |  |  |  |
| No | 101 | 0 | 0.38* |
| Yes | 867 | 13 |  |
| Comorbidities | 319 | 9 | **0.017\*** |
| Cardiovascular | 13 | 0 | 0.81 |
| Immunodeficiency | 27 | 2 | **0.03** |
| Malignancy | 11 | 1 | 0.21 |
| Neuromuscular | 24 | 1 | **0.046** |
| Hypertension | 49 | 4 | **0.005** |
| CKD | 3 | 1 | **< 0.0001** |
| Lung disease | 31 | 1 | 0.47 |
| DM | 16 | 1 | 0.28 |
| Duration of the<br>disease (years) |  |  | 0.357* |
| 0-10 | 444 | 5 |  |
| 11-20 | 522 | 3 |  |
| 21-30 | 47 | 1 |  |
| 31-40 | 14 | 0 |  |
| >40 | 2 | 0 |  |
| MS type |  |  |  |
| RR | 894 | 12 | 0.73 |
| Progressive | 102 | 2 |  |
| Other | 130 | 1 |  |
| DMT |  |  |  |
| Low efficacy | 216 | 5 | 0.59* |
| Medium efficacy | 293 | 3 |  |
| High efficacy | 175 | 2 |  |
| other | 183 | 2 |  |

## DISCUSSION

The intricate relationship between COVID-19 and people with multiple sclerosis (pwMS) has emerged as a compelling area of investigation during the global pandemic. Our study, based on the expansive dataset from the MS Global Data Sharing Initiative, provided by PhysioNet, provides vital information on this intersection. A key finding from our study is the significant role of comorbidities in determining the outcomes of COVID-19 in pwMS. Specifically, neuromuscular disorders, hypertension, CKD, and immunodeficiencies emerged as critical factors. Neuromuscular disorders can compromise respiratory function, leading to an increased susceptibility to respiratory complications from COVID-19, a trend observed in the wider population ^9^. Hypertension has been consistently identified as a risk factor for severe outcomes of COVID-19, potentially due to associated cardiovascular complications that can be exacerbated by the virus ^10^. Immunodeficiencies, whether inherent or acquired, can attenuate the body’s ability to mount an effective response against SARS-CoV-2, leading to extended and more severe disease courses ^11^.

Our study offers a fresh perspective on the relationship between disease-modifying therapies (DMT) and the severity of COVID-19 in pwMS. Contrary to the initial fears of the MS community, our analysis suggests that high-efficacy DMTs do not exacerbate the severity of COVID-19. This observation is consistent with several other studies and alleviates concerns about the use of DMT in MS patients during the ongoing pandemic ^12, 13^. A prospective analysis of 40 MS patients also showed that the type of DMT did not significantly influence the severity of COVID-19 ^14^. It underscores the notion that while DMTs modify the immune response, they do not necessarily increase susceptibility to severe infections, including those caused by SARS-CoV-2. One hypothesis suggests that immunomodulators may be inversely correlated with COVID-19-related mortality due to decreased inflammation and cytokine storm; however, more studies are needed to validate this ^15^. Similarly, prior use of steroids as a treatment was not correlated with worse outcomes.

The type of MS, whether it was relapsing-remitting or primary progressive, did not emerge as a significant determinant of the severity of COVID-19 in our cohort. This suggests that the inherent pathophysiology of the specific MS type may be less influential in determining the outcomes of COVID-19 compared to other factors such as age, duration of the disease, or associated comorbidities. Despite this, few studies have shown otherwise. Studies have demonstrated that people with progressive MS are more likely to be hospitalized than those with RRMS^16, 17^. Furthermore, a Swedish registry cohort analysis found that those with progressive MS are more likely to have a severe infection ^18^. *Januel et al*. also demonstrated that individuals with primary progressive MS are more likely to develop a severe COVID-19 infection, and anti-CD20 therapy may also be associated with the worst outcomes in those with RRMS ^19^. However, it is worth noting that the role of MS type in infectious disease outcomes remains a debated topic, warranting further investigation.

Our study, although comprehensive, has several limitations. Reliance on self-reported or clinically reported data introduces potential biases. The granularity of the data set did not allow us to dive deeper into the severity or control of comorbid conditions, which can significantly influence the outcomes. Additionally, the data set lacked adequate information on individual EDSS scores, and only a small proportion of the reported sample size tested positive for COVID-19. The lack of a contemporaneous non-MS control group restricts direct comparisons. The data’s temporal limitation, with the collection tool being decommissioned in early 2022, also means that the long-term implications of COVID-19 on this cohort remain an enigma. While our study sheds light on the immediate relationship between MS, its therapeutic management, and COVID-19, the long-term landscape remains shrouded in uncertainty. Longitudinal studies focusing on long COVID or post-acute sequelae of SARS-CoV-2 infection in pwMS are crucial. One such analysis in pwMS after COVID-19 infection did not demonstrate an increase in long-term disease activity ^20^. In addition, mechanistic studies to elucidate the underlying biology of the observed associations could not only clarify these relationships but also pave the way for therapeutic advancements in the management of viral infections in pwMS.

## CONCLUSIONS

Using a comprehensive data set from the MS Global Data Sharing Initiative provided by PhysioNet, we were able to dissect various factors that influence the outcomes of COVID-19 in people with multiple sclerosis (pwMS). A key takeaway from our research is the pronounced impact of comorbidities such as neuromuscular disorders, hypertension, CKD, and immunodeficiencies in determining the severity of COVID-19 outcomes in pwMS. This underscores the importance of comprehensive health assessment that extends beyond the primary disease condition to include associated comorbidities for this patient population. Type of MS, duration of the disease, and high-efficacy DMT did not significantly influence the outcomes of COVID-19. Although our study provides a robust perspective, it also opens avenues for future research, especially in exploring the mechanisms by which DMT and different types of MS may or may not influence the severity of infectious diseases such as COVID-19.

## Data Availability

All data produced in the present study are available upon reasonable request to the authors

## Acknowledgements

We would like to thank all people with MS and clinicians for their time invested in providing the data within the context of the GDSI. Additionally, we sincerely thank the original curators of this database for making this important work open-access for analysis and dissemination into the greater academic community.

